# Household cooking fuels, symptoms compatible with pneumonia and inequalities in care seeking among children under five in the Republic of the Congo: a nationally representative survey analysis

**DOI:** 10.64898/2026.10.26.26366480

**Authors:** Esthel Lee Presley Bemba, A. P. Oko, Franck Hardain Okemba Okombi, Axhel Ronan Malonga Miafouna, Armel Batchi-Bouyou, Darius Eryx Mbou Essie, Marguyvonne Hornelie Loksley Ginnella, Rany Eba-Gakabe

## Abstract

**Objectives:** To examine the association between household cooking fuels and symptoms compatible with pneumonia, and socioeconomic inequalities in care seeking, among children under five in the Republic of the Congo.

**Methods:** Secondary analysis of the 2014–2015 Multiple Indicator Cluster Survey, the most recent national survey with available microdata on respiratory symptoms (9,159 children aged 0– 59 months), using survey-weighted logistic regression.

**Results:** The weighted prevalence of caretaker-reported cough with fast or difficult breathing due to a chest problem was 5.9% (95% CI 5.3–6.6). Compared with electricity or gas, cooking with solid fuels (adjusted odds ratio [aOR] 2.23, 1.34–3.73) or kerosene (aOR 2.63, 1.38–5.01) was associated with higher odds of symptoms, independently of wealth, education and residence (E-values 3.9 and 4.7). Only 28.2% (23.1–33.9) of children with symptoms were taken to a health facility or provider, from 17.8% in the poorest to 61.1% in the richest quintile (aOR 4.10, 1.96– 8.57 for the three richest versus the two poorest quintiles).

**Conclusions:** Poorer Congolese children combine greater exposure to polluting cooking fuels and less access to care. Clean cooking policies should be combined with removal of financial barriers to care for sick children.

## Background

Lower respiratory infections, mainly pneumonia, remain the leading infectious cause of death among children under five worldwide, and the burden is concentrated in sub-Saharan Africa [1–3]. Most deaths could be prevented by vaccination, reduction of exposure to known risk factors, and prompt assessment and antibiotic treatment by a trained provider [3,4]. Established risk factors include young age, low birth weight, undernutrition, lack of exclusive breastfeeding, crowding and exposure to household air pollution [4].

Household air pollution from cooking with polluting fuels caused an estimated 309,000 deaths among children under five in 2021 [5]. A meta-analysis estimated that exposure to unprocessed solid fuels increases the risk of pneumonia in young children by a factor of about 1.8 [6], a randomised trial of improved cookstoves in Guatemala reduced severe pneumonia [7], and recent multi-country analyses of household surveys in sub-Saharan Africa continue to report associations between polluting fuels or smoke exposure and respiratory symptoms in children [8,9]. Kerosene, often promoted as a transition fuel, emits fine particles and other pollutants and is classified by the World Health Organization as a polluting fuel [5,10]. Beyond exposure, survival depends on whether caregivers take the child to a qualified provider; care seeking for suspected pneumonia remains low and inequitable in many sub-Saharan African countries [11].

In the Republic of the Congo, a lower-middle-income country of Central Africa where most of the population lives in Brazzaville and Pointe-Noire, the pneumococcal conjugate vaccine was introduced into routine immunisation in 2012 [12]. Yet most children live in households that cook with wood or charcoal [13], and the national survey report presents only descriptive tabulations of respiratory symptoms and care seeking [13]. The joint role of household energy and socioeconomic position, and the specific contribution of kerosene, which is widely used in Congolese cities, have not been examined.

Using the 2014–2015 Multiple Indicator Cluster Survey (MICS), the most recent national survey with individual-level data on these indicators, this study aimed to estimate the prevalence of symptoms compatible with pneumonia among children under five, to examine their association with household cooking fuels and other child and household characteristics, and to identify factors associated with care seeking from a health facility or provider among children with symptoms. We hypothesised that younger age and small size at birth (H1), and exposure to polluting cooking fuels and crowding (H2), would be associated with higher odds of symptoms, and that poverty, low maternal education and rural residence would be associated with lower odds of care seeking (H3).

## Methods

### Study design and data source

We conducted a secondary analysis of cross-sectional data from the fifth round of MICS in the Republic of the Congo, implemented by the Institut National de la Statistique with technical and financial support from UNICEF [13]. Fieldwork took place from November 2014 to February 2015. The survey is catalogued under the reference COG_2014_MICS_v01_M in the World Bank Microdata Library and the International Household Survey Network catalogue. The study is reported following the STROBE guidelines [16] (Checklist S1).

### Choice of data source

We reviewed the nationally representative household surveys conducted in the Republic of the Congo since 2010. The 2011–2012 Demographic and Health Survey [17] predates MICS5, the 2021 national nutrition survey did not include the respiratory module [14], and the 2025 Demographic and Health Survey has published only key indicators, without microdata, based on 74 weighted children with symptoms of acute respiratory infection [15]. MICS5 is therefore the most recent source allowing an individual-level analysis of these associations, and provides a baseline for later comparison with the 2025 data.

### Sampling

The survey used a two-stage stratified cluster sample designed to provide estimates at national level, for urban and rural areas and for the 12 departments. The sampling frame was the cartography of the 2007 General Census of Population and Housing. Within each of 15 strata defined by department and area of residence, enumeration areas were selected with probability proportional to size, and a fixed number of households was selected systematically in each enumeration area. Of 13,275 sampled households, 12,811 were interviewed (99.6% of occupied households). The mothers or caretakers of all children under five in interviewed households were eligible for the under-five questionnaire; interviews were completed for 9,169 of 9,271 eligible children (98.9%), in 530 clusters.

### Study population and missing data

The household, women’s and under-five data files were merged using the cluster, household and line numbers. We included all children aged 0–59 months with a completed under-five questionnaire. Ten children whose caretakers did not know or did not report whether the child had a cough, fast or difficult breathing, or the location of the breathing problem were excluded, leaving 9,159 children for the analysis of symptoms. The analysis of care seeking was restricted to the 624 children with symptoms. Missing data on explanatory variables were rare (cooking fuel 22 children, including one household that did not cook; maternal education 2), and the models were fitted on complete cases (9,135 children in the main model).

### Outcomes

Symptoms compatible with pneumonia were defined according to the standard MICS indicator [13,18]: the child had an illness with a cough in the two weeks preceding the interview (question CA7), accompanied by short, rapid breaths or difficulty breathing (CA8) that was due to a problem in the chest or to both a problem in the chest and a blocked or running nose (CA9).

Children whose breathing problem was due only to a blocked or running nose were classified as not having symptoms. This indicator is a caretaker-reported syndrome, not a clinical or radiological diagnosis of pneumonia [18].

Care seeking was defined as advice or treatment sought from a health facility or provider (CA11): public hospital, health centre, health post, community health worker or other public medical source, private hospital or clinic, private physician, mobile clinic or other private medical source. Private pharmacies, shops, relatives or friends and traditional practitioners were not counted, consistent with the MICS indicator. A sensitivity analysis counted private pharmacies as providers.

### Explanatory variables

Child characteristics were age, grouped as 0–11, 12–23 and 24–59 months (0–23 and 24–59 months in the care-seeking analysis because of the smaller sample), and sex. Size at birth as perceived by the mother (smaller than average or very small, versus average or larger) was available from the women’s questionnaire only for the last live birth in the two years preceding the survey; it was linked to the child through the mother’s line number and the child’s month and year of birth.

Household environmental characteristics were the main cooking fuel, grouped as clean (electricity, liquefied petroleum gas), kerosene, and solid (charcoal, wood, straw, sawdust); the place of cooking, recorded only for households not using electricity, gas or kerosene, which we combined with fuel type into four categories (non-solid fuel; solid fuel used outdoors, in a separate building, or inside the house); and crowding, defined as more than three household members per room used for sleeping.

Socioeconomic characteristics were the mother’s or caretaker’s highest level of education, the household wealth index quintile provided with the dataset (constructed by principal component analysis of household assets and dwelling characteristics), and area of residence. In the regression models, education was grouped as none or primary versus secondary or higher, and wealth as the two poorest versus the three richest quintiles; models with all five quintiles are presented as sensitivity analyses.

Variables used in sensitivity analyses were stunting (height-for-age z-score below −2 according to the WHO growth standards, excluding flagged measurements), a natural earth or sand floor as a proxy for housing quality, current breastfeeding, and receipt of three doses of pneumococcal conjugate vaccine according to the vaccination card or the caretaker’s report. Breastfeeding and vaccination were collected only for children under three years. The survey did not collect information on household tobacco smoke exposure or ventilation.

### Statistical analysis

All estimates used the children’s sample weights and accounted for stratification (15 strata) and clustering (530 enumeration areas) by Taylor series linearisation, giving 515 design degrees of freedom (530 clusters minus 15 strata) for Wald F tests and confidence intervals. Proportions are presented with 95% confidence intervals computed on the logit scale. Associations between each explanatory variable and each outcome were tested with design-adjusted Wald F tests.

We fitted survey-weighted logistic regression models with design-based standard errors. Model 1 examined the association between symptoms and child age, sex, cooking fuel, crowding, maternal education, wealth and area of residence in all children. All candidate variables were entered simultaneously, without stepwise selection, because the aim was to estimate adjusted associations rather than to build a prediction model. Model 2 examined the association between care seeking and child age, sex, maternal education, wealth and area of residence among children with symptoms; crude odds ratios are presented alongside adjusted odds ratios (aOR).

Model calibration was assessed with the Archer–Lemeshow goodness-of-fit test for survey data [19], which compares weighted observed and predicted outcomes across deciles of predicted probability with a design-based F test, and with a calibration plot (Figure S1); discrimination was summarised by the weighted area under the receiver operating characteristic curve (AUC).

Prespecified sensitivity analyses of Model 1 (i) added size at birth among children aged 0–23 months, (ii) replaced fuel type with the combined fuel and cooking-location variable, (iii) added stunting and floor material, (iv) added department fixed effects, and (v) among children aged 12– 35 months, added stunting, current breastfeeding and pneumococcal vaccination; the last model was compared with the same model without these variables in the same subsample. Sensitivity analyses of Model 2 counted private pharmacies as providers, used all five wealth quintiles, and removed wealth from the model to examine its role in the associations with education and residence. To assess how strong unmeasured confounding would have to be to explain the associations with cooking fuel, we computed E-values for the adjusted odds ratios and their lower confidence limits [20], treating odds ratios as approximations of risk ratios because the outcome was rare (prevalence below 10% in every fuel category).

Analyses were performed in Python 3 (pandas, NumPy, SciPy) with survey estimators implemented from standard linearisation formulas. All prevalence estimates, both main models and the goodness-of-fit tests were independently reproduced in R 4 with the survey package [21] (svydesign, svyciprop, svyglm, regTermTest), with identical point estimates; confidence limits differed by at most 0.1 percentage point for proportions and 0.02 for odds ratios, owing to different reference distributions. We report effect sizes with 95% confidence intervals and interpret them according to their magnitude and precision rather than p-value thresholds. The analysis code is publicly available (see Data availability statement).

## Results

### Characteristics of the study population

Of the 9,159 children included, 50.9% were boys, 20.6% were aged 0–11 months and 18.6% were aged 12–23 months (weighted percentages, Table 1). Most children lived in urban areas (62.5%) and in households cooking mainly with solid fuels (66.9%; wood 39.6%, charcoal 26.9%); 22.8% lived in households using electricity or gas and 10.3% in households using kerosene, 92.5% of whom lived in urban areas. Among children in households using solid fuels, cooking took place outdoors for 62.9%.

**Table 1.** Characteristics of children aged 0–59 months and prevalence of symptoms compatible with pneumonia, Republic of the Congo, MICS 2014–2015 (n = 9,159).

| Characteristic | n (unweighted) | Weighted % | Prevalence of symptoms, % (95% CI) | p |
| --- | --- | --- | --- | --- |
| Child age |  |  |  | 0.041 |
| 0–11 months | 1,889 | 20.6 | 5.4 (4.3–6.8) |  |
| 12–23 months | 1,770 | 18.6 | 7.8 (6.3–9.8) |  |
| 24–59 months | 5,500 | 60.8 | 5.5 (4.6–6.5) |  |
| Sex |  |  |  | 0.154 |
| Female | 4,523 | 49.1 | 5.4 (4.6–6.3) |  |
| Male | 4,636 | 50.9 | 6.4 (5.4–7.5) |  |
| Size at birth (perceived)<sup>a</sup> |  |  |  | 0.121 |
| Average or larger | 2,821 | 90.7 | 6.0 (5.0–7.1) |  |
| Smaller than average / very small | 338 | 9.3 | 8.7 (5.6–13.2) |  |
| Main cooking fuel |  |  |  | <0.001 |
| Clean (electricity, LPG) | 1,008 | 22.8 | 2.5 (1.6–3.8) |  |
| Kerosene | 488 | 10.3 | 6.4 (4.4–9.2) |  |
| Solid (charcoal, wood, other biomass) | 7,641 | 66.9 | 7.0 (6.2–7.9) |  |
| Cooking fuel and location<sup>b</sup> |  |  |  | <0.001 |
| Non-solid fuel | 1,496 | 33.2 | 3.7 (2.8–4.8) |  |
| Solid fuel, separate building | 2,905 | 16.2 | 7.7 (6.4–9.3) |  |
| Solid fuel, inside the house | 1,175 | 8.5 | 8.1 (5.6–11.4) |  |
| Solid fuel, outdoors | 3,544 | 42.2 | 6.6 (5.6–7.8) |  |
| Household crowding |  |  |  | 0.660 |
| ≤3 persons per sleeping room | 5,536 | 59.3 | 6.0 (5.3–6.9) |  |
| >3 persons per sleeping room | 3,623 | 40.7 | 5.7 (4.7–6.9) |  |
| Mother's education |  |  |  | 0.111 |
| None | 1,094 | 8.3 | 6.5 (4.7–8.8) |  |
| Primary | 3,148 | 25.2 | 7.1 (6.0–8.4) |  |
| Lower secondary | 3,865 | 46.0 | 5.5 (4.6–6.6) |  |
| Upper secondary or higher | 1,050 | 20.4 | 5.2 (3.9–7.0) |  |
| Wealth index quintile |  |  |  | <0.001 |
| Poorest | 4,185 | 24.0 | 8.1 (7.0–9.4) |  |
| Second | 2,355 | 22.0 | 7.7 (6.0–9.8) |  |
| Middle | 1,196 | 20.4 | 5.8 (4.3–7.8) |  |
| Fourth | 807 | 18.1 | 3.7 (2.5–5.7) |  |
| Richest | 616 | 15.5 | 2.6 (1.5–4.4) |  |
| Area of residence |  |  |  | <0.001 |
| Rural | 6,448 | 37.5 | 8.0 (7.2–9.0) |  |
| Urban | 2,711 | 62.5 | 4.6 (3.8–5.6) |  |

CI, confidence interval. p, design-adjusted Wald F test. Percentages are weighted and computed among children with non-missing data; n are unweighted counts. Missing data: cooking fuel 22 (including one household that did not cook); cooking fuel and location 39; maternal education 2.
<sup>a</sup> Available only for the last live birth in the two years preceding the survey (n = 3,159).
<sup>b</sup> Place of cooking was recorded only for households not using electricity, gas or kerosene.

### Prevalence of symptoms compatible with pneumonia

Overall, 624 children had symptoms compatible with pneumonia in the two weeks preceding the interview, a weighted prevalence of 5.9% (95% CI 5.3–6.6). Prevalence was higher in rural (8.0%, 7.2–9.0) than in urban areas (4.6%, 3.8–5.6) and ranged from 2.5% (1.6–3.7) in Pointe-Noire and 3.2% (2.0–5.2) in Plateaux to 9.2% (6.6–12.7) in Sangha and 10.2% (8.4–12.4) in Likouala (Figure 1).

**Figure 1.**
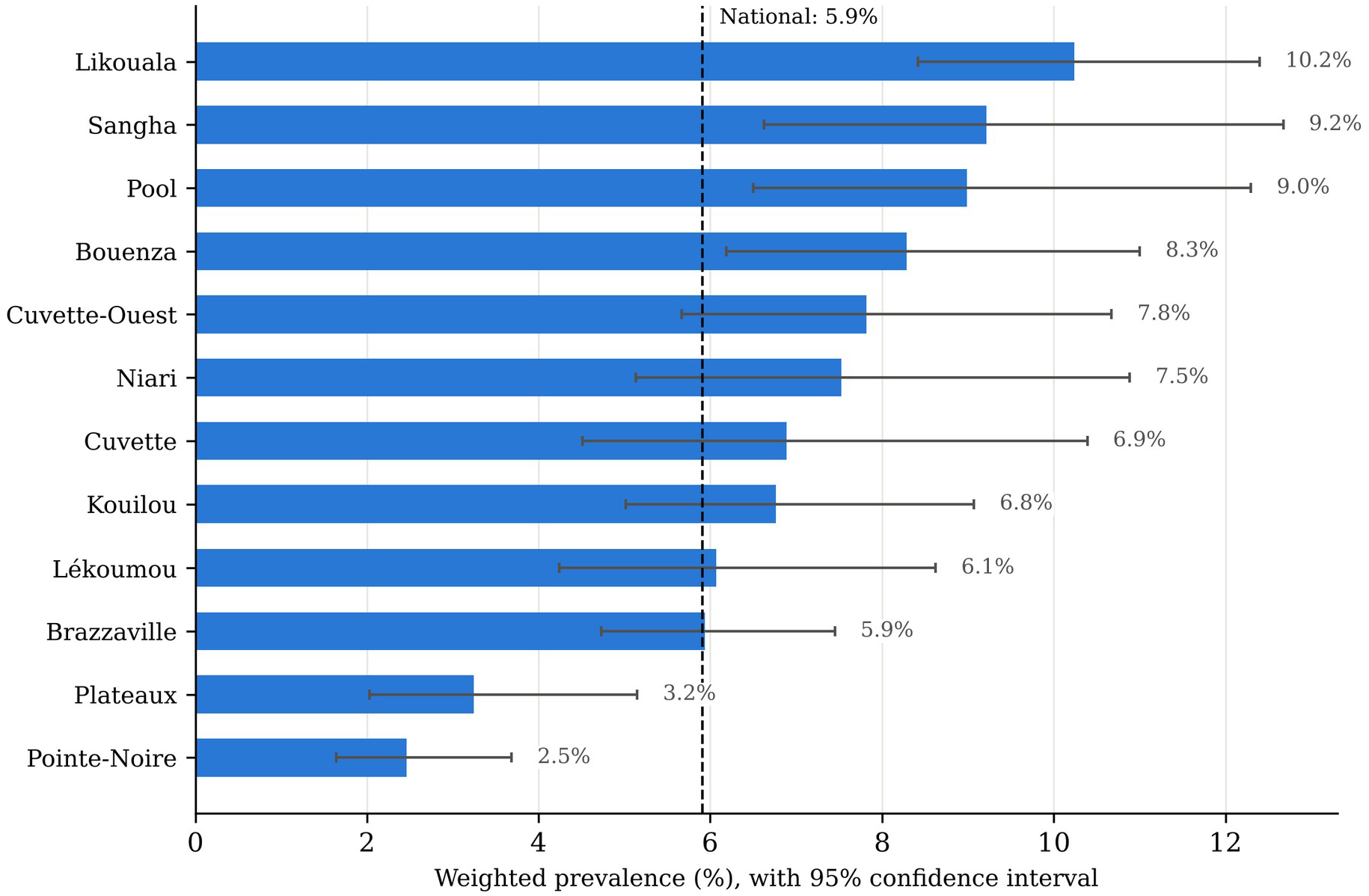
Prevalence of symptoms compatible with pneumonia among children aged 0–59 months, by department, Republic of the Congo, MICS 2014–2015. Bars show weighted prevalence and error bars 95% confidence intervals; the dashed line shows the national prevalence.

In bivariate analysis, prevalence was highest at 12–23 months (7.8%) and was higher among children in households using solid fuels (7.0%) or kerosene (6.4%) than among those using clean fuels (2.5%). It decreased with household wealth, from 8.1% in the poorest to 2.6% in the richest quintile. Children reported to be small at birth had a higher prevalence than other children (8.7% versus 6.0%), with overlapping confidence intervals. Crowding was not associated with symptoms (Table 1).

### Factors associated with symptoms compatible with pneumonia

In Model 1 (Table 2), children in households cooking with solid fuels (aOR 2.23, 95% CI 1.34– 3.73) or kerosene (aOR 2.63, 1.38–5.01) had higher odds of symptoms than children in households using clean fuels. Children aged 12–23 months had higher odds than those aged 24– 59 months (aOR 1.48, 1.08–2.03), whereas infants aged 0–11 months did not (aOR 0.98, 0.72– 1.33). After adjustment, wealth (aOR 1.40, 0.85–2.31 for the two poorest quintiles), area of residence, maternal education and crowding were not clearly associated with symptoms (confidence intervals including 1). The model showed no evidence of poor calibration (Archer– Lemeshow F = 1.19, p = 0.29; Figure S1); its discrimination was modest (AUC 0.63).

**Table 2.** Factors associated with symptoms compatible with pneumonia among children aged 0–59 months, survey-weighted multivariable logistic regression (Model 1).

| Variable | Category | aOR | 95% CI | p |
| --- | --- | --- | --- | --- |
| Child age | 0–11 months | 0.98 | 0.72–1.33 | 0.885 |
|  | 12–23 months | 1.48 | 1.08–2.03 | 0.014 |
|  | 24–59 months | 1.00 | Reference |  |
| Sex | Male | 1.19 | 0.93–1.53 | 0.163 |
|  | Female | 1.00 | Reference |  |
| Main cooking fuel | Kerosene | 2.63 | 1.38–5.01 | 0.003 |
|  | Solid (charcoal, wood, other biomass) | 2.23 | 1.34–3.73 | 0.002 |
|  | Clean (electricity, LPG) | 1.00 | Reference |  |
| Household crowding | >3 persons per sleeping room | 0.86 | 0.67–1.11 | 0.250 |
|  | ≤3 persons per sleeping room | 1.00 | Reference |  |
| Mother's education | None or primary | 0.93 | 0.74–1.17 | 0.533 |
|  | Secondary or higher | 1.00 | Reference |  |
| Wealth | Poorest two quintiles | 1.40 | 0.85–2.31 | 0.183 |
|  | Middle to richest quintiles | 1.00 | Reference |  |
| Area of residence | Rural | 1.22 | 0.81–1.86 | 0.341 |
|  | Urban | 1.00 | Reference |  |

aOR, adjusted odds ratio; CI, confidence interval. n = 9,135 children (624 with symptoms); 24 children with missing data on cooking fuel or maternal education were excluded. Archer–Lemeshow goodness-of-fit test: $F(10, 515) = 1.19$ , $p = 0.29$ . Area under the ROC curve 0.63.

The associations with cooking fuel were robust in sensitivity analyses (Table S2). They were little changed after adjustment for stunting and floor material, for department, and, among children aged 12–35 months, for breastfeeding and pneumococcal vaccination, although confidence intervals were wide in this subsample (n = 3,133). The E-values were 3.89 (2.01 for the lower confidence limit) for solid fuels and 4.70 (2.10) for kerosene: an unmeasured confounder would need to be associated with both cooking fuel and symptoms by a risk ratio of about 3.9 for solid fuels and 4.7 for kerosene, beyond the measured covariates, to fully explain the observed associations.

### Care seeking for children with symptoms

Advice or treatment was sought from any source for 252 of the 624 children with symptoms (weighted 40.8%, 95% CI 35.0–46.8), and from a health facility or provider for 148 (28.2%, 23.1–33.9). The most frequent sources were health centres (11.9%) and public hospitals (10.7%), followed by relatives or friends (5.5%) and private pharmacies (3.4%); private medical providers and community health workers were rarely used (Table S1). Overall, 183 children with symptoms (28.1%, 23.2–33.5) received an antibiotic.

Care seeking increased with household wealth, from 17.8% in the poorest to 61.1% in the richest quintile, and was more frequent in urban than in rural areas (34.3% versus 22.3%) and among children whose mother had secondary or higher education (32.8% versus 21.1%) (Table 3). In Model 2, only wealth remained associated with care seeking (aOR 4.10, 1.96–8.57 for the three richest versus the two poorest quintiles). The crude associations with maternal education (OR 1.83, 1.06–3.14) and urban residence (OR 1.81, 1.06–3.08) disappeared after adjustment for wealth. Wealth and residence overlapped strongly: 91% of rural children with symptoms belonged to the two poorest quintiles, against 31% of urban children. With all five quintiles, the odds of care seeking increased across the wealth gradient (aOR 4.70, 6.73 and 13.39 for the middle, fourth and richest quintiles versus the poorest), with wide confidence intervals because only 121 children with symptoms belonged to the three richest quintiles (Table S3). Results were similar when private pharmacies were counted as providers (aOR for wealth 2.98, 1.32–6.76).

**Table 3.** Care seeking from a health facility or provider among children with symptoms compatible with pneumonia: proportions, crude and adjusted odds ratios (Model 2), n = 624.

| Characteristic | n (unweighted) | Care sought, % (95% CI) | Crude OR (95% CI) | aOR (95% CI) | p |
| --- | --- | --- | --- | --- | --- |
| Child age |  |  |  |  |  |
| 0–23 months | 278 | 32.3 (24.7–40.9) | 1.42 (0.80–2.53) | 1.53 (0.85–2.77) | 0.158 |
| 24–59 months | 346 | 25.1 (18.2–33.4) | 1.00 (reference) | 1.00 (reference) |  |
| Sex |  |  |  |  |  |
| Female | 310 | 29.0 (22.1–37.0) | 1.00 (reference) | 1.00 (reference) |  |
| Male | 314 | 27.5 (20.9–35.3) | 0.93 (0.56–1.53) | 0.91 (0.56–1.49) | 0.711 |
| Mother's education |  |  |  |  |  |
| None or primary | 335 | 21.1 (15.1–28.6) | 1.00 (reference) | 1.00 (reference) |  |
| Secondary or higher | 289 | 32.8 (25.4–41.1) | 1.83 (1.06–3.14) | 0.90 (0.50–1.63) | 0.737 |
| Wealth |  |  |  |  |  |
| Poorest two quintiles | 503 | 18.9 (14.1–24.8) | 1.00 (reference) | 1.00 (reference) |  |
| Middle to richest quintiles | 121 | 43.2 (32.8–54.3) | 3.27 (1.86–5.74) | 4.10 (1.96–8.57) | <0.001 |
| Area of residence |  |  |  |  |  |
| Rural | 500 | 22.3 (17.4–28.2) | 1.00 (reference) | 1.00 (reference) |  |
| Urban | 124 | 34.3 (25.3–44.6) | 1.81 (1.06–3.08) | 0.81 (0.38–1.71) | 0.573 |

OR, odds ratio; aOR, adjusted odds ratio; CI, confidence interval. Model 2 includes all variables shown. Archer–Lemeshow goodness-of-fit test: $F = 0.35$ , $p = 0.97$ ; area under the ROC curve 0.67. Only 121 children with symptoms belonged to the three richest quintiles; estimates by quintile are shown in Table S3.

## Discussion

In this nationally representative survey, about one in seventeen children under five in the Republic of the Congo had symptoms compatible with pneumonia in the two weeks before the interview, with higher prevalence in rural areas and large differences between departments.

Living in a household that cooked with solid fuels or kerosene was associated with more than twice the odds of symptoms, independently of wealth and residence, with consistent estimates across sensitivity analyses, although some subgroup estimates were imprecise. Fewer than three in ten children with symptoms were taken to a health facility or provider, and household wealth was the only factor independently associated with care seeking. Beyond the descriptive national report [13], this analysis adds adjusted associations, the specific role of kerosene and the joint examination of exposure and access to care.

### Household air pollution and symptoms compatible with pneumonia

The association with solid fuel use (aOR 2.2) is consistent with the pooled estimate of about 1.8 from a meta-analysis of household air pollution and childhood pneumonia [6] and with recent analyses of household surveys in sub-Saharan Africa [8,9]. The association with kerosene, of similar magnitude, adds to the evidence that kerosene is not a clean alternative to biomass [5,10]. Cooking outdoors or in a separate building was not clearly associated with lower odds of symptoms among solid fuel users; young children are often carried by the person cooking, and smoke from neighbouring households also contributes to exposure, so the place of cooking is a crude proxy for personal exposure. Residual confounding cannot be excluded, but the stability of the estimates across adjustment sets and E-values of about four argue against a large bias.

The age pattern, with the highest prevalence at 12–23 months rather than in infancy, did not support our first hypothesis. Reported symptoms depend on the caretaker’s recognition of breathing difficulty, which may be lower in young infants, and breastfeeding may protect infants during the first months of life [4]. The association with size at birth was weak and imprecise, but this analysis was limited to recent births and had low power.

### Socioeconomic inequalities in care seeking

Care seeking from a health facility or provider (28.2%) was low, as in many countries of sub-Saharan Africa [11], and strongly patterned by wealth, with a monotonic gradient across quintiles. The associations of care seeking with maternal education and urban residence were explained statistically by wealth. Because rural children with symptoms almost all belong to the two poorest quintiles, the effects of wealth and residence cannot be fully separated in this sample. These findings are compatible with financial barriers to care, such as consultation fees and the cost of medicines and transport, but the data do not allow us to distinguish them from other correlates of wealth, such as distance to facilities, perceived quality of care or caretakers’ recognition of danger signs. The high proportion of children for whom no care was sought, or who were treated by relatives, pharmacies or traditional practitioners, means that many children with possible pneumonia were not assessed by a trained provider.

### A double disadvantage for poorer children

Taken together, the results describe a double disadvantage. Children in poorer households were more likely to live with polluting cooking fuels, which were associated with symptoms compatible with pneumonia, and, when they had symptoms, they were less likely to be taken to a health facility. The attenuation of the association between wealth and symptoms after adjustment for cooking fuel is consistent with fuel use lying on the pathway between poverty and respiratory symptoms, but cross-sectional regression adjustment does not establish mediation, and wealth and fuel use may also share other unmeasured determinants.

### Implications for policy

The 2025 Demographic and Health Survey key indicators (about 1% of children with symptoms of acute respiratory infection; advice or treatment sought for 50.1%) are not directly comparable with ours: the care-seeking indicator includes shops and drug vendors (32.9%, 27.3–38.9, with a similar definition in our data), it rests on 74 weighted cases, and fieldwork took place in a different season [15]. Our findings support expanding access to clean cooking, including liquefied petroleum gas and electricity rather than kerosene, together with free care for sick children under five and community case management of pneumonia in underserved rural areas.

### Strengths and limitations

Strengths include a large nationally representative sample, standardised indicators, full account of the survey design with independent reproduction in a second software, and multiple sensitivity analyses. The study also has limitations. First, symptoms compatible with pneumonia were reported by caretakers and have limited validity against clinical diagnosis; many children with these symptoms do not have pneumonia, and some children with pneumonia are missed [18]. Our results therefore concern reported respiratory symptoms, not confirmed pneumonia. Second, the cross-sectional design does not allow causal inference, and symptoms and care seeking are subject to recall bias. Third, exposure to household air pollution was measured indirectly through the main cooking fuel and place of cooking, without information on stove type, ventilation, ambient air pollution or tobacco smoke. Fourth, some covariates were available only for subsets of children, and few children with symptoms belonged to the richest quintiles. Finally, the data are about ten years old; changes in household energy, user-fee policies or vaccination coverage since 2015 may have modified both the levels and the associations reported here. The lower prevalence of reported symptoms in 2025 [15], although not directly comparable, is a further reason to re-examine these associations with the 2025 microdata.

## Conclusions

In the Republic of the Congo, children living in households that cook with solid fuels or kerosene had higher odds of symptoms compatible with pneumonia, and fewer than one in three children with these symptoms was taken to a health facility or provider, with a steep wealth gradient.

Poorer children thus combine greater exposure to a modifiable environmental risk and lower access to care. Policies that accelerate access to clean cooking and remove financial barriers to care for sick children should be evaluated together, and updated national data will be needed to monitor progress.

## Supporting information

STROBE checklist

## Data Availability

The data analysed in this study are from the 2014-2015 Republic of the Congo Multiple Indicator Cluster Survey (MICS5) and are available from UNICEF (https://mics.unicef.org/surveys) and the World Bank Microdata Library (reference COG_2014_MICS_v01_M), subject to registration and acceptance of the terms of use of the data providers. All results produced in the present study are contained in the manuscript and its supplementary files. The analysis code is available from the corresponding author upon reasonable request.

## Acknowledgements

We thank UNICEF and the Institut National de la Statistique of the Republic of the Congo for making the MICS 2014–2015 data available, and the households who participated in the survey. A large language model was used to assist with editing the manuscript; the authors performed and verified all analyses and take full responsibility for the content.

## Ethics statement

This study is a secondary analysis of anonymised data from the 2014–2015 Republic of the Congo Multiple Indicator Cluster Survey (MICS5), conducted by the Institut National de la Statistique with the support of UNICEF following the standard MICS protocols, under which informed consent is obtained from respondents and from the mothers or caretakers of children before interview [13]. The authors used de-identified microdata (World Bank Microdata Library reference COG_2014_MICS_v01_M), had no access to personal identifiers and did not contact any participant. The study was approved by the Comité d’Éthique de la Recherche en Sciences de la Santé (CERSSA), Republic of the Congo, in September 2025 (reference no. 138-25/MESRSIT/DGRST/CERSSA/-25).

## Data availability statement

The data analysed in this study are from the 2014–2015 Republic of the Congo Multiple Indicator Cluster Survey (MICS5) and are available from UNICEF (https://mics.unicef.org/surveys) and the World Bank Microdata Library (reference COG_2014_MICS_v01_M), subject to registration and acceptance of the terms of use of the data providers. All results produced in the present study are contained in the manuscript and its supplementary files. The analysis code is available from the corresponding author on reasonable request.

## Supporting information

**Checklist S1.** STROBE checklist for cross-sectional studies.

Table S1. Sources of advice or treatment for children with symptoms compatible with pneumonia (n = 624).

Table S2. Sensitivity analyses for symptoms compatible with pneumonia.

Table S3. Sensitivity analyses for care seeking.

Figure S1. Calibration of Model 1: weighted observed versus mean predicted prevalence of symptoms by decile of predicted probability.

