## Supplementary Tables S1-S3 and Figure S1 for "Household cooking fuels, symptoms compatible with pneumonia and inequalities in care seeking among children under five in the Republic of the Congo: a nationally representative survey analysis"

**Table S1. Sources of advice or treatment for children with symptoms compatible with pneumonia (n = 624).**

| **Indicator** | n (unweighted) | Weighted % (95% CI) |
|---|---|---|
| Advice or treatment sought from any source | 252 | 40.8 (35.0–46.8) |
| Health facility or provider (excluding pharmacy) | 148 | 28.2 (23.1–33.9) |
| Health facility or provider, or private pharmacy | 160 | 31.2 (25.7–37.3) |
| Received an antibiotic | 183 | 28.1 (23.2–33.5) |
| Source: Public hospital | 52 | 10.7 (7.4–15.2) |
| Source: Health centre | 64 | 11.9 (8.3–16.8) |
| Source: Health post | 13 | 2.6 (0.9–7.2) |
| Source: Community health worker | 2 | 0.1 (0.0–0.4) |
| Source: Other public medical | 1 | 0.1 (0.0–0.7) |
| Source: Private hospital | 3 | 0.5 (0.2–1.6) |
| Source: Private physician | 3 | 0.9 (0.2–3.5) |
| Source: Private pharmacy | 15 | 3.4 (1.7–6.9) |
| Source: Private or mobile clinic | 9 | 1.1 (0.5–2.6) |
| Source: Other private medical | 2 | 0.4 (0.1–1.5) |
| Source: Relative or friend | 54 | 5.5 (3.7–8.1) |
| Source: Shop | 17 | 1.6 (1.0–2.8) |
| Source: Traditional practitioner | 17 | 1.9 (1.1–3.5) |
| Source: Other | 9 | 0.8 (0.4–1.8) |

Several sources could be reported for the same child; sources do not add up to the total.

**Table S2 (a). Sensitivity analyses of Model 1: adjusted odds ratios for cooking fuel (reference: electricity or gas).**

| **Model** | n | With symptoms | Solid fuels aOR (95% CI) | Kerosene aOR (95% CI) | Archer–Lemeshow p | Added variables, aOR (95% CI) |
|---|---|---|---|---|---|---|
| Main model (Model 1) | 9,135 | 624 | 2.23 (1.34–3.73) | 2.63 (1.38–5.01) | 0.29 | |
| + stunting and floor material | 8,611 | 583 | 2.46 (1.41–4.29) | 2.99 (1.54–5.80) | 0.53 | Stunted (height-for-age z < −2) 1.22 (0.90–1.64); Earth or sand floor 1.62 (0.98–2.68) |
| + department fixed effects | 9,135 | 624 | 2.10 (1.25–3.53) | 2.31 (1.21–4.39) | 0.86 | |
| Children 12–35 months, base model | 3,133 | 248 | 1.84 (0.84–4.04) | 1.95 (0.78–4.84) | 0.04 | |
| Children 12–35 months, + stunting, breastfeeding, PCV3 | 3,133 | 248 | 1.88 (0.85–4.14) | 2.03 (0.81–5.06) | 0.22 | Stunted (height-for-age z < −2) 1.31 (0.85–2.02); Currently breastfed 1.06 (0.68–1.66); Received 3 doses of PCV 1.37 (0.92–2.05) |

All models also adjust for age, sex, crowding, maternal education, wealth and area of residence. PCV3, three doses of pneumococcal conjugate vaccine. Breastfeeding and vaccination were collected only for children under 36 months.

**Table S2 (b). Model 1b: children aged 0–23 months with information on size at birth (n = 3,055, 234 with symptoms).**

| **Variable** | Category | aOR | 95% CI | p |
|---|---|---|---|---|
| Child age | 0–11 months | 0.72 | 0.49–1.05 | 0.087 |
| | 12–23 months | 1.00 | Reference | |
| Sex | Male | 1.21 | 0.80–1.81 | 0.361 |
| | Female | 1.00 | Reference | |
| Main cooking fuel | Kerosene | 1.85 | 0.55–6.25 | 0.324 |
| | Solid (charcoal, wood, other biomass) | 2.70 | 0.93–7.82 | 0.068 |
| | Clean (electricity, LPG) | 1.00 | Reference | |
| Household crowding | >3 persons per sleeping room | 1.05 | 0.71–1.53 | 0.819 |
| | ≤3 persons per sleeping room | 1.00 | Reference | |
| Mother's education | None or primary | 0.89 | 0.64–1.24 | 0.497 |
| | Secondary or higher | 1.00 | Reference | |
| Wealth | Poorest two quintiles | 1.27 | 0.66–2.43 | 0.471 |
| | Middle to richest quintiles | 1.00 | Reference | |
| Area of residence | Rural | 1.49 | 0.83–2.68 | 0.179 |
| | Urban | 1.00 | Reference | |
| Size at birth (perceived) | Smaller than average / very small | 1.28 | 0.75–2.16 | 0.362 |
| | Average or larger | 1.00 | Reference | |

aOR, adjusted odds ratio.

**Table S2 (c). Model 1c: cooking fuel combined with place of cooking (n = 9,118).**

| **Variable** | Category | aOR | 95% CI | p |
|---|---|---|---|---|
| Child age | 0–11 months | 0.98 | 0.71–1.34 | 0.893 |
| | 12–23 months | 1.48 | 1.08–2.04 | 0.015 |
| | 24–59 months | 1.00 | Reference | |
| Sex | Male | 1.21 | 0.94–1.55 | 0.134 |
| | Female | 1.00 | Reference | |
| Household crowding | >3 persons per sleeping room | 0.89 | 0.69–1.16 | 0.392 |
| | ≤3 persons per sleeping room | 1.00 | Reference | |
| Mother's education | None or primary | 0.94 | 0.74–1.19 | 0.618 |
| | Secondary or higher | 1.00 | Reference | |
| Wealth | Poorest two quintiles | 1.48 | 0.87–2.50 | 0.147 |
| | Middle to richest quintiles | 1.00 | Reference | |

| | | | | |
|---|---|---|---|---|
| Area of residence | Rural | 1.22 | 0.78–1.89 | 0.386 |
| | Urban | 1.00 | Reference | |
| Cooking fuel and location | Solid fuel, separate building | 1.36 | 0.86–2.16 | 0.190 |
| | Solid fuel, inside the house | 1.56 | 0.89–2.74 | 0.119 |
| | Solid fuel, outdoors | 1.44 | 0.98–2.11 | 0.066 |
| | Non-solid fuel | 1.00 | Reference | |

aOR, adjusted odds ratio.

**Table S3 (a). Sensitivity analyses of Model 2 (care seeking from a health facility or provider, n = 624).**

| **Model and variable** | aOR (95% CI) | p |
|---|---|---|
| Model 2 with five wealth quintiles (reference: poorest) | | |
| Age 0–23 months | 1.49 (0.84–2.63) | 0.172 |
| Male | 0.98 (0.61–1.58) | 0.940 |
| Mother secondary or higher | 0.82 (0.44–1.53) | 0.534 |
| Urban | 0.61 (0.29–1.32) | 0.211 |
| Second quintile | 1.55 (0.76–3.19) | 0.228 |
| Middle quintile | 4.70 (1.80–12.24) | 0.002 |
| Fourth quintile | 6.73 (2.03–22.28) | 0.002 |
| Richest quintile | 13.39 (3.18–56.35) | <0.001 |
| Model 2 without wealth | | |
| Age 0–23 months | 1.47 (0.81–2.64) | 0.203 |
| Male | 0.92 (0.56–1.50) | 0.728 |
| Mother secondary or higher | 1.50 (0.85–2.62) | 0.159 |
| Urban | 1.55 (0.89–2.71) | 0.121 |

Reference categories: age 24–59 months, female, mother with no or primary education, rural residence.

**Table S3 (b). Model 2 with private pharmacies counted as providers (n = 624, 160 with care sought).**

| **Variable** | Category | aOR | 95% CI | p |
|---|---|---|---|---|
| Child age | 0–23 months | 1.39 | 0.81–2.39 | 0.226 |
| | 24–59 months | 1.00 | Reference | |
| Sex | Male | 0.99 | 0.60–1.62 | 0.959 |
| | Female | 1.00 | Reference | |
| Mother's education | Secondary or higher | 1.19 | 0.66–2.14 | 0.555 |
| | None or primary | 1.00 | Reference | |
| Wealth | Middle to richest quintiles | 2.98 | 1.32–6.76 | 0.009 |
| | Poorest two quintiles | 1.00 | Reference | |
| Area of residence | Urban | 1.02 | 0.49–2.10 | 0.962 |
| | Rural | 1.00 | Reference | |

aOR, adjusted odds ratio.

**Figure S1. Calibration of Model 1: weighted observed versus mean predicted prevalence of symptoms compatible with pneumonia by decile of predicted probability. The dashed line indicates perfect calibration.**

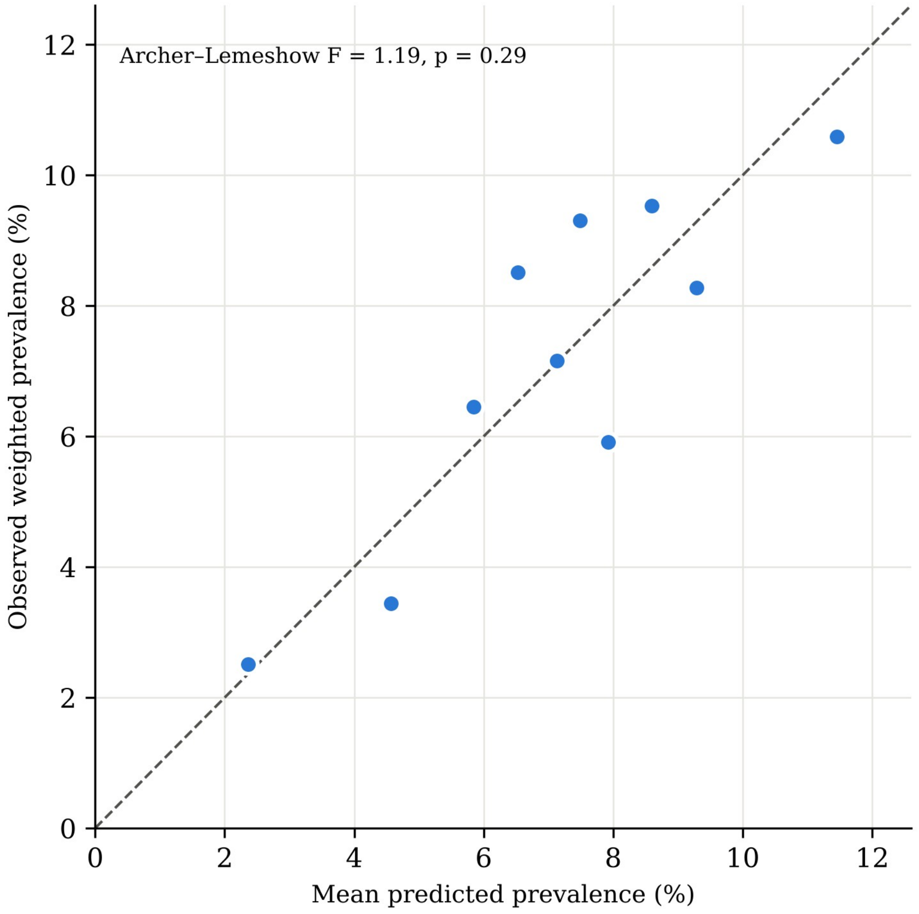
