## Supplementary material for "Household cooking fuels, symptoms compatible with pneumonia and inequalities in care seeking among children under five in the Republic of the Congo: a nationally representative survey analysis": STROBE checklist

**S1 Checklist. STROBE statement: checklist of items for cross-sectional studies**

| Item | No. | Recommendation | Location in manuscript |
|---|---|---|---|
| Title and abstract | 1 | Indicate the study design in the title or abstract; provide an informative and balanced abstract | Title ("a nationally representative survey analysis"); Abstract (cross-sectional 2014–2015 Multiple Indicator Cluster Survey) |
| Background/rationale | 2 | Explain the scientific background and rationale | Background, paragraphs 1–3 |
| Objectives | 3 | State specific objectives, including any prespecified hypotheses | Background, last paragraph (H1–H3) |
| Study design | 4 | Present key elements of study design early in the paper | Methods: Study design and data source |
| Setting | 5 | Describe the setting, locations, and relevant dates | Methods: Study design and data source; Choice of data source; Sampling |
| Participants | 6 | Give the eligibility criteria, and the sources and methods of selection of participants | Methods: Sampling; Study population and missing data |
| Variables | 7 | Clearly define all outcomes, exposures, predictors, potential confounders | Methods: Outcomes; Explanatory variables |
| Data sources/measurement | 8 | Give sources of data and details of methods of assessment | Methods: Outcomes; Explanatory variables (questionnaire items cited) |
| Bias | 9 | Describe any efforts to address potential sources of bias | Methods: Statistical analysis (survey weights, design, E-values); Discussion: Strengths and limitations |
| Study size | 10 | Explain how the study size was arrived at | Methods: Sampling; Study population and missing data |
| Quantitative variables | 11 | Explain how quantitative variables were handled | Methods: Explanatory variables (age groups, crowding threshold, grouping of education and wealth) |
| Statistical methods | 12a-e | Describe statistical methods, subgroups, missing data, sampling strategy, sensitivity analyses | Methods: Statistical analysis; Study population and missing data |
| Participants | 13 | Report numbers of | Methods: Sampling; |

| | | | |
|---|---|---|---|
| | | individuals at each stage of study | Study population and missing data |
| Descriptive data | 14 | Give characteristics of study participants; indicate number with missing data | Results: Characteristics of the study population; Table 1; Table 2 footnote |
| Outcome data | 15 | Report numbers of outcome events or summary measures | Results: Prevalence; Care seeking; Tables 1 and 3; Figure 1 |
| Main results | 16 | Give unadjusted and adjusted estimates and their precision | Tables 1–3; Tables S2 and S3 |
| Other analyses | 17 | Report other analyses done (subgroups, interactions, sensitivity analyses) | Results; Figure S1; Tables S2 and S3 |
| Key results | 18 | Summarise key results with reference to study objectives | Discussion, paragraph 1 |
| Limitations | 19 | Discuss limitations, sources of potential bias or imprecision | Discussion: Strengths and limitations |
| Interpretation | 20 | Give a cautious overall interpretation of results | Discussion (four subsections); Conclusions |
| Generalisability | 21 | Discuss the generalisability (external validity) | Discussion: Strengths and limitations |
| Funding | 22 | Give the source of funding and the role of the funders | Funding statement |
